# Comparing family history indicators and polygenic scores in depressive disorder

**DOI:** 10.64898/2026.01.19.26343941

**Authors:** Emma Pruin, Yuri Milaneschi, Meike Bartels, Pietro Bassani, Brenda Penninx, Wouter J. Peyrot

## Abstract

**Background:** Genetic liability of depressive disorder can be captured by psychopathology in relatives (family history). Various methods summarize family history in a single score, differing in included information as well as underlying model. We systematically compared the performance of family history indicators, including promising new indicators based on the liability threshold model, in predicting depressive disorder.

**Methods:** We calculated selected family history indicators for depression (dichotomous, proportion, novel genetically-informed method PAFGRS) in 1339 participants of the Netherlands Study of Depression and Anxiety (*N_case_*= 1086). Polygenic scores were computed from the most recent GWAS for major depression. We assessed correlations between genetic liability indicators, as well as their prediction of lifetime depressive disorder diagnosis.

**Results:** Correlations of family history indicators with each other were high (*r* = 0.71 - 0.99), and much lower with the PGS (*r* = 0.15). There was a suggested increase in predictive accuracy for more elaborately computed scores, ranging from proportion (*AUC* = 0.66, *OR* = 2.26, *95%CI* = 1.88-2.71) to PAFGRS (*AUC* = 0.70, *OR* =17.06, *95%CI* = 9.46 - 30.77). The best-performing family history indicator and the PGS were independently associated with depressive disorder (PAFGRS: *OR* = 15.17, *95%CI* = 8.36-27.51, *p* = 3.59×10^-19^; PGS: *OR* = 1.30, *95%CI* = 1.12-1.50, *p* = 0.0004).

**Conclusions:** Our analysis shows that more elaborate family history indicators, including family size, prevalence, heritability and based on genetic theory, would be preferrable over simpler methods. Family history and PGS were complementary in prediction, showing the added value of including both in future studies.

## Introduction

Individual vulnerability to being diagnosed with a psychiatric disorder, such as depressive disorder, is partially explained by genetic variation. In capturing this genetic liability, common variation in the human genome has recently taken center stage, with a spotlight on the development and improvement of polygenic scores (PGS). However, PGS explain only a small portion of the variance in complex traits, and many practical and ethical issues concerning their use in clinical practice are unresolved (Murray 2020, Adeyemo 2021). In contrast, assessment of genetic liability through family history has long been established as a valuable part of medical risk assessment (Pyeritz 2012, Guttmacher 2004, Yoon 2002). Family history has been linked to liability for disease, as represented by risk of diagnosis or severity-related clinical characteristics, in psychiatric disorders (Hardt & Franke 2007), including common mood and anxiety disorders (Havinga 2017, McLoughlin 2008, Nierenberg 2007, Hettema 2001, Sullivan 2000). Nevertheless, the potential of family history data for prediction of psychiatric disorders has not been fully explored.

For disorder prediction, it is important to optimally assess and summarize family history data. The commonly used way of enquiring about family history is to determine the presence or absence of a disorder in the (extended) family (e.g. Mars 2022). More detailed, elaborate indicators can be presumed to be more informative and powerful. An example is the count of affected relatives, which can be extended with additional family-specific information, such as family size (Ali 2021, Milne 2009), age of relatives (van Sprang 2022), age of onset in relatives (Pedersen 2022), or cohabitation (Kendler 2021). More elaborate indicators also make use of disorder-specific information, such as prevalence (Ruderfer 2010, van Sprang 2022) and heritability (So 2011, Hujoel 2020), or cross-trait genetic correlations (Krebs 2025). Indicators further differ in the ways they are calculated, including analytical algorithms of varying complexity (Verdoux 1996, So 2011), maximum likelihood estimation (Hujoel 2020, Pedersen 2022), and step-by-step heuristics (Kendler 2021). In consequence, heterogeneous and sample-specific methodologies prohibit systematic comparison of existing results.

A comprehensive and flexible approach to summarize family history has recently been developed based on the liability threshold model (Falconer 1965, So et al. 2011). One application, the Pearson-Aitkens Family Genetic Risk Score (PAFGRS) models genetic liabilities based on family- and disease specific information, including genetic covariances, using a computationally cheap analytical approach (Krebs 2025). PAFGRS showed better prediction of depression than other elaborate approaches, but was not compared to simple dichotomous or proportional indicators. Additionally, its application and performance was assessed in a registry-based study, and has not yet been validated in a clinical cohort assessed with structured interviews.

Here, we leveraged data from 1339 participants of the Netherlands Study of Depression and Anxiety (NESDA) (Penninx 2021), to compare the performance of different genetic liability indicators in predicting lifetime depressive disorder diagnoses. We calculated simple and elaborate family history indicators from detailed interviews on family history of depression and anxiety in first degree relatives. Our main aim was to compare different types of family history indicators in predicting depressive disorder. Furthermore, we benchmarked the prediction of the family history indicator against that of a PGS for major depression derived from the latest genome-wide association study meta-analysis (Adams 2025).

## Methods

### NESDA Sample

The Netherlands Study of Anxiety and Depression (NESDA) is a longitudinal cohort study aimed at investigating the course and consequences of anxiety and depressive disorders. It is described in detail elsewhere (Penninx et al. 2021). Between 2003 and 2007, participants with depressive and anxiety disorders and healthy controls were recruited from the community, primary care, and secondary care settings. Exclusion criteria were a) having a severe psychiatric disorder other than those of interest in NESDA, e.g. psychotic disorders, obsessive-compulsive disorder, bipolar disorder or addiction disorder, and b) not being fluent in Dutch. Follow-ups have taken place at intervals of 1-3 years, resulting in 6 completed waves in 2023. The Ethical Review Board of the VU University Medical Centre (Medisch Ethische Toetsingscommissie Vumc) gave ethical approval for the study protocol of NESDA. All participants signed informed consent.

After exclusion of closely related participants (full siblings), the final sample of the current study consisted of 1339 individuals with available family history and genotype data. 1086 of these participants reported a DSM-based diagnosis of depressive disorder during psychiatric interviews at one of the NESDA assessments, for 253 no depressive disorder was ascertained.

### Lifetime diagnosis with depressive disorder

Lifetime diagnosis of depressive disorder (MDD or dysthymia) as defined in the DSM-IV (Diagnostic and Statistical Manual of Mental Disorders) was assessed using the Composite Interview Diagnostic Instrument (version 2.1, WHO), administered by trained research staff at baseline. By the same method, diagnostic status was updated at the 3rd, 4th, 5^th^, and 6th follow-up. Screened participants without any lifetime diagnosis of depressive disorder, anxiety disorder, or bipolar disorder were assigned to the control group.

### Family history assessment

Family history was assessed at Wave 6 using the Family Tree Inventory interview (Fyer & Weissman 1999) in Dutch. First, each participant was asked general questions about the composition of their family. Separate questions were used to ascertain the number of biological brothers and sisters, excluding half-, step-, foster- and adoptive siblings. Family size was calculated as the number of siblings (excluding the participant) plus two parents. Next, the existing first-degree relatives were considered one-by-one regarding basic descriptives and their likely diagnostic status.

If the presence of psychological problems was suggested for a given relative, the interviewer moved on to the specific disorder questions for depression, then anxiety. If the participant then indicated relevant symptoms, follow-up questions pertaining to whether they had accessed professional help, medication, inpatient treatment and ECT were asked, with the purpose of increasing the reliability of self-report family diagnoses.

We compared two ways of coding the interview – here referred to as short versus extended assessment. The short assessment was based on a single core question of whether the relative had ever had an episode of depression or anxiety. For the extended assessment, relatives were classified as affected only if there was an indication that they had undergone any treatments for depression or anxiety. An illustration with the original questions can be found in **Figure S1**, precise instructions in **Supplemental Methods**. Direct comparison of the two assessments showed only small differences in sample sizes and prevalences (see **Table S1**).

The diagnosis of the family members was coded as ‘missing’ under the following conditions: (a) when the answer to the core question was missing or “don’t know”, and (b) when the follow up questions did not confirm diagnosis (extended assessment only). When disorder status of a family member was missing, this family member was omitted from the data, but the participant themselves was retained. We also assessed the effect of including relatives with missing data as controls instead, here referred to as imputation. This did not substantially improve sample size (**Table S2** ).

Given the lack of demonstrated benefit of extended assessment or imputation, the main results presented here are based exclusively on the short assessment. We conducted secondary association analyses on extended assessment and imputation.

### Family history indicators

We describe below how different family history indicators were calculated in this study. Figure 1 displays the input parameters considered by each of the indicators, with reference to the example of depression.

**Figure 1:**
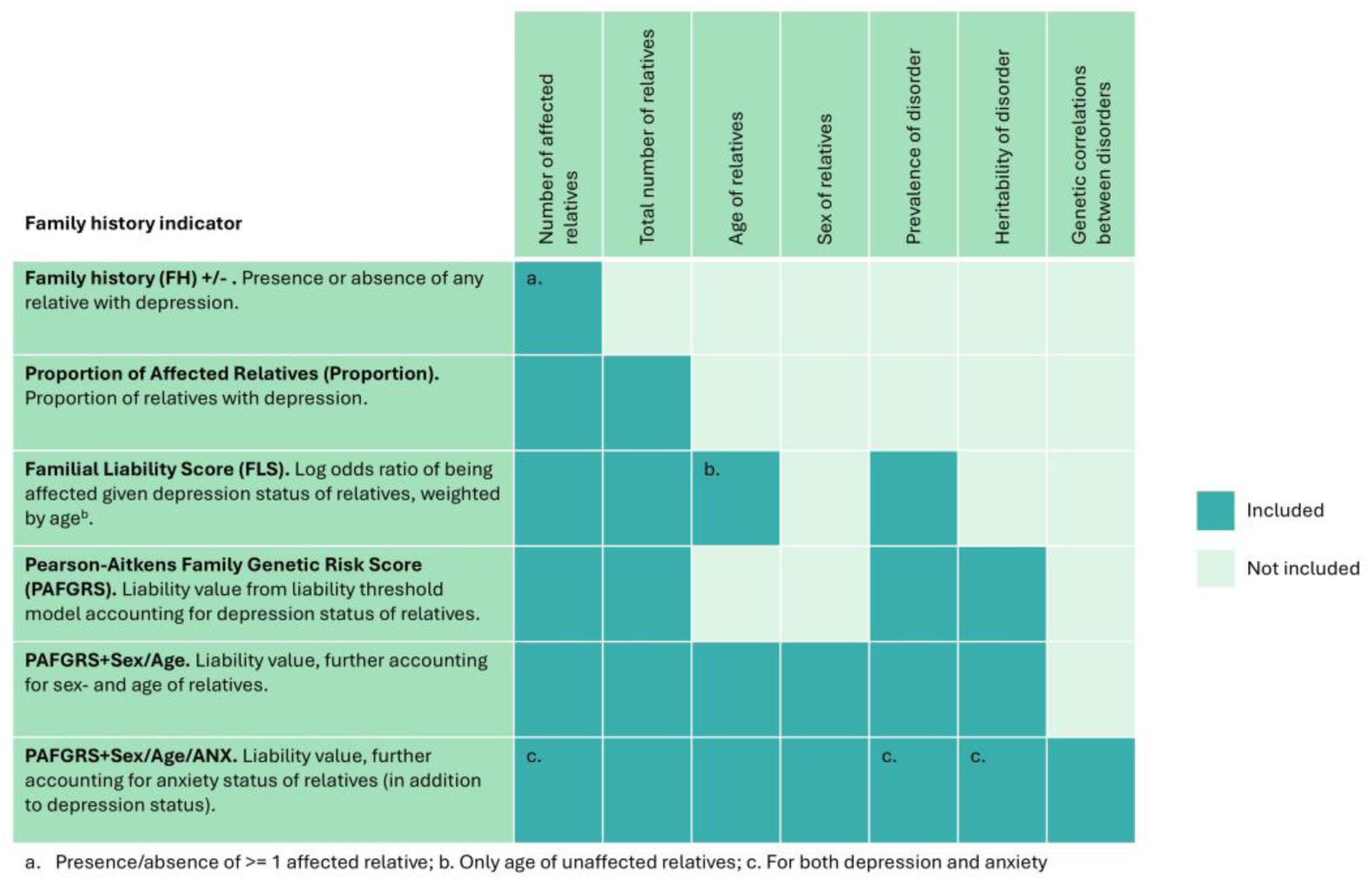
Overview of family history indicators and their required input.

### FH+/-

Family history (FH)+/- is a simple dichotomous score, indicating absence versus presence of any relatives with depression. Thus, if one or more relatives had received a diagnosis with depression, this indicator would be set to 1 (= FH+), else to 0 (= FH-).

### Proportion

Proportion was calculated by dividing the number of relatives with a depression diagnosis by the total number of relatives. Thus, this indicator ranges between 0 and 1, with the number of possible levels depending on family size.

### FLS

The Familial Liability Score (FLS) is a continuous indicator based on a combination of likelihood ratios per relative (Verdoux et al. 1996). This indicator accounts for the age of unaffected relatives, following the rationale that there is a higher certainty of low liability in individuals who have remained unaffected for a longer time. We followed the FLS algorithm as previously applied in NESDA by van Sprang et al. (2020) to depression, with the same parameters for age-of-onset range (10-65), lifetime prevalence (K) in those with affected relatives (K = 0.40), and lifetime prevalence in those without affected relatives (K = 0.094).

### PAFGRS

Pearson-Aitkens Family Genetic Risk Score (PAFGRS) is based on the liability threshold model: Genetic liability for depression can be modeled in each relative based on their disease status and lifetime population prevalence of the disease (Krebs et al. 2025). Covariances and variances of genetic liabilities in the family follow from relatedness and heritability (h²). Using normal distribution theory, participants’ genetic liability was adjusted based on relatives’ liabilities to obtain a risk score. We used the R package PAFGRS (v0.1.0, Krebs et al. 2024) with input of twin heritability h² = 0.35 (Polderman et al. 2015) and lifetime prevalence K = 0.19 for depression (de Graaf et al. 2012).

### PAFGRS+Sex/Age

PAFGRS+Sex/Age is an extension of the PAFGRS indicator that allows for a personalized prevalence estimate, based on each relative’s sex and age. Sex- and age-specific prevalences were drawn from a continuous curve built from published age bin prevalences, scaled for the population prevalence of the Netherlands (McGrath et al. 2023, de Graaf et al. 2012) (**Figure S2**). In relatives without age information, we used the sex-specific average prevalence (de Graaf et al. 2012). Contrary to FLS, PAFGRS+Sex/Age uses age information for both affected and unaffected relatives. The related intuition is that older age in unaffected relatives corresponds to lower liability; and younger age in affected relatives corresponds to higher liability.

### PAFGRS+Sex/Age/ANX

PAFGRS+Sex/Age/ANX further extends the PAFGRS indicator, in addition to sex- and age-specific risk, with information on comorbid anxiety. As such, the participants liability for depression was increased by presence of anxiety disorder in a relative, modelled with h² = 0.40 (Polderman et al. 2015) and K = 0.20 (de Graaf et al. 2012) for anxiety. The effect of family history of anxiety on the liability for depression was scaled by the genetic correlation of the two disorders (rg = 0.68, Grotzinger et al. 2022).

### PGS

Imputed genotype data was available for 2541 NESDA participants of European Ancestry. PGS were built for these individuals based on summary statistics for major depression (Adams et al. 2025) in European Ancestry individuals (excluding 23andMe, and the Dutch NESDA and Rotterdam study cohorts; approximately 410374 cases and 1584648 controls) using the 1000 Genomes European ancestry reference (phase 3; Auton et al. 2015). PGS were generated using plink2 (Purcell et al. 2007) and PRS-CS (v1.1.0). PRS-CS is a Bayesian method that weighs single-nucleotide polymorphisms by a continuous shrinkage factor to account for LD and performs well compared to other methods (Ge et al. 2019). PGS were standardized to mean = 0 and variance = 1. Further details on imputation and preprocessing of the genotype data can be found in **Supplemental Methods**.

### Statistical analyses

Correlations between the indicators were calculated using Pearson’s correlation; secondary analyses were based on Spearman’s rho correlation given the dichotomous nature of the FH+/-variable.

To compare predictive performance of the indicators, we conducted logistic regression of depressive disorder on all indicators separately. To assess the combined predictive performance of PAFGRS+Sex/Age/ANX and PGS, both were included in the same regression model as predictors. Regression coefficients and p-values were inspected. For the main regression models, the receiver operator curve (ROC) -- a metric of the true positive rate and the false positive rate of a prediction model – was summarized as area under the curve (AUC) using the R package pROC (version X, Robin et al. 2011).

Additionally, to visualize incremental risk by higher score, we investigated prediction of depressive disorder separately per quintile of PAFGRS+Sex/Age/ANX and PGS using a logistic regression model with quintile as a predictor, consequently transforming coefficients and standard errors to odds ratios and confidence intervals.

## Results

### Descriptives of the NESDA sample and family history

Of the 1339 NESDA participants with available family history data and genotype data, 1086 had been diagnosed with depressive disorder at some point in their lives, and 53 had never been diagnosed with a psychiatric disorder (**Table 1**). Participants were predominantly female (54.15% in controls, 65.47% in cases), with a mean age of 42 years. The median family size was four 1st degree relatives, i.e. two parents and two siblings, with 50% of participants reporting between three and five 1st degree relatives (one and three siblings). Across indicators, the mean familial risk for depression was larger in cases than controls, with similar variance in the two groups.

**Table 1:** Sample characteristics. Dep.: Depressive disorder; SD: Standard deviation; IQR: Inter-quartile range; FH+/-: Family history of depression present/absent; FH+: Family history of depression present; FLS: Familial Liability Score; PAFGRS: Pearson-Aitkens Family Genetic Risk Score; ANX: Anxiety; PGS: Polygenic Score. *PGS was standardized to mean = 0 and sd = 1 in full sample.

|  | Controls | Dep. Disorder Cases |
| --- | --- | --- |
| **Sample characteristics** |  |  |
| N | 253 | 1086 |
| Age (Mean [SD]) | 43.00 (14.24) | 42.46 (12.60) |
| Sex (% female) | 54.15 | 65.47 |
| Number of 1st degree relatives (Median [IQR; Range]) | 4 (3-5; 2-14) | 4 (3-5; 2-17) |
| **Family history indicators for depression** |  |  |
| FH+/- (% FH +) | 44.27 | 68.6 |
| Proportion (Mean [SD]) | 0.16 (0.24) | 0.32 (0.35) |
| FLS (Mean [SD]) | -0.11 (0.63) | 0.25 (0.72) |
| PAFGRS (Mean [SD]) | -0.03 (0.24) | 0.14 (0.28) |
| PAFGRS+Sex/Age (Mean [SD]) | -0.03 (0.24) | 0.15 (0.29) |
| PAFGRS+Sex/Age/ANX (Mean [SD]) | -0.10 (0.24) | 0.10 (0.28) |
| PGS (Mean [SD]) | -0.31 (1.02)* | 0.05 (1.00)* |

Distributions of both family history indicators and PGS show a distinction between depressive disorder cases and controls, with higher levels in cases (**Figure 2**). Peaks in family history indicator density plots correspond to the number of affected individuals in families of different sizes (apart from the dichotomous FH+/-). By definition, PGS are normally distributed.

**Figure 2:**
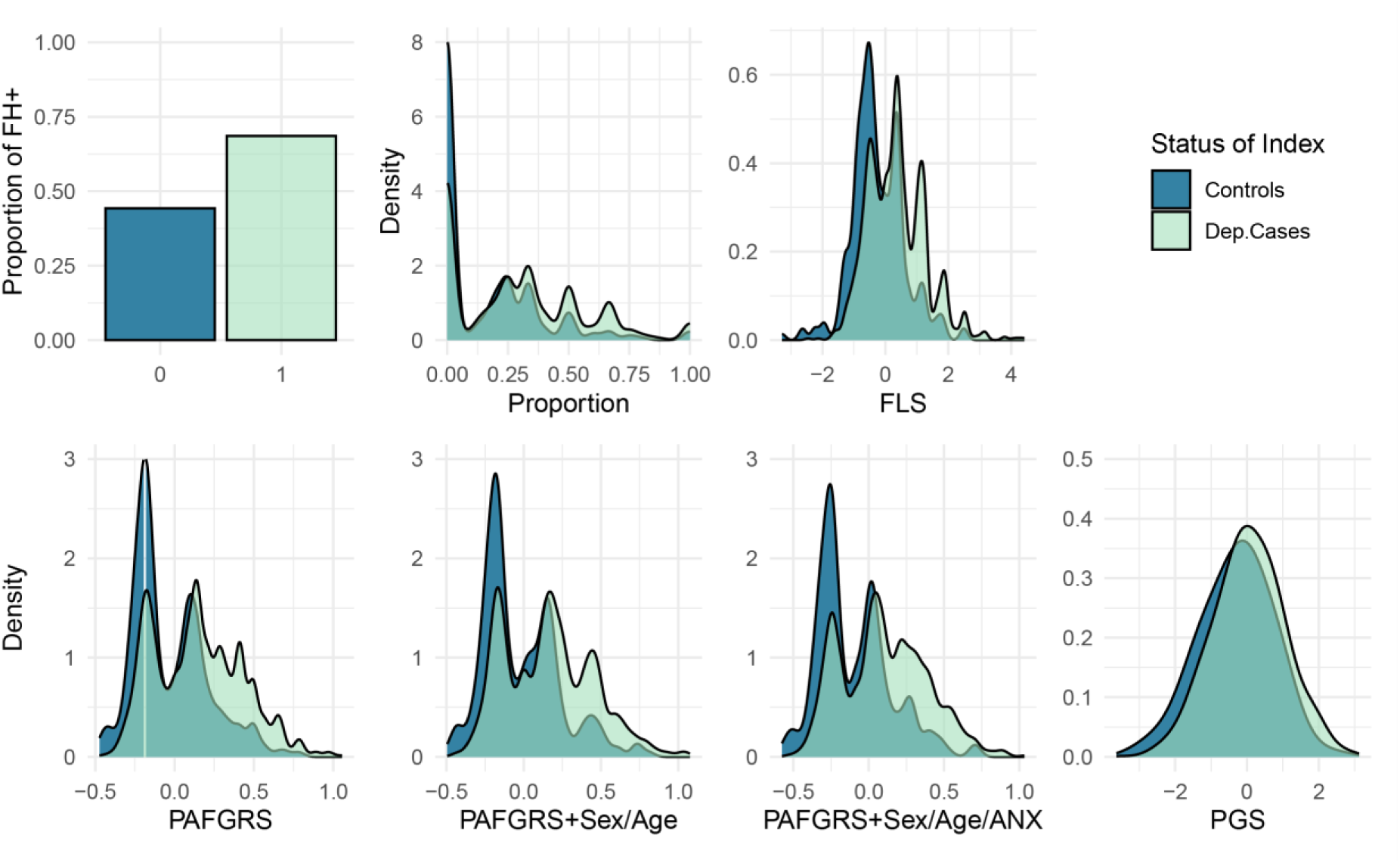
Density plots of family history indicators by depressive disorder. Dep.: Depressive disorder; FH+: Family history of depression present; FLS: Familial Liability Score; PAFGRS: Pearson-Aitkens Family Genetic Risk Score; ANX: Anxiety; PGS: Polygenic Score.

### Relationship between family history indicators and PGS

The family history indicators were highly correlated but not identical (**Figure 3**). Correlations between the continuous family history indicators Proportion, FLS, PAFGRS, PAFGRS+Sex/Age, PAFGRS+Sex/Age/ANX were particularly high, ranging from r = 0.885 to r = 0.991. Specifically, the three PAFGRS scores were highly correlated to each other (0.971-0.991), the FLS score (0.955-0.973), and the proportion score (0.898-0.935). FH+/- was slightly less correlated to the other indicators (r = 0.706 - 0.782), due to ignoring detailed family history information and as a statistical consequence of contrasting a dichotomous indicator to a non-dichotomous indicator. In contrast, correlations between the PGS and the family history indicators were small (r = 0.115 - 0.167). The presented results are based on Pearson correlation estimations; Spearman’s rank correlations show a similar pattern (**Figure S3**).

**Figure 3:**
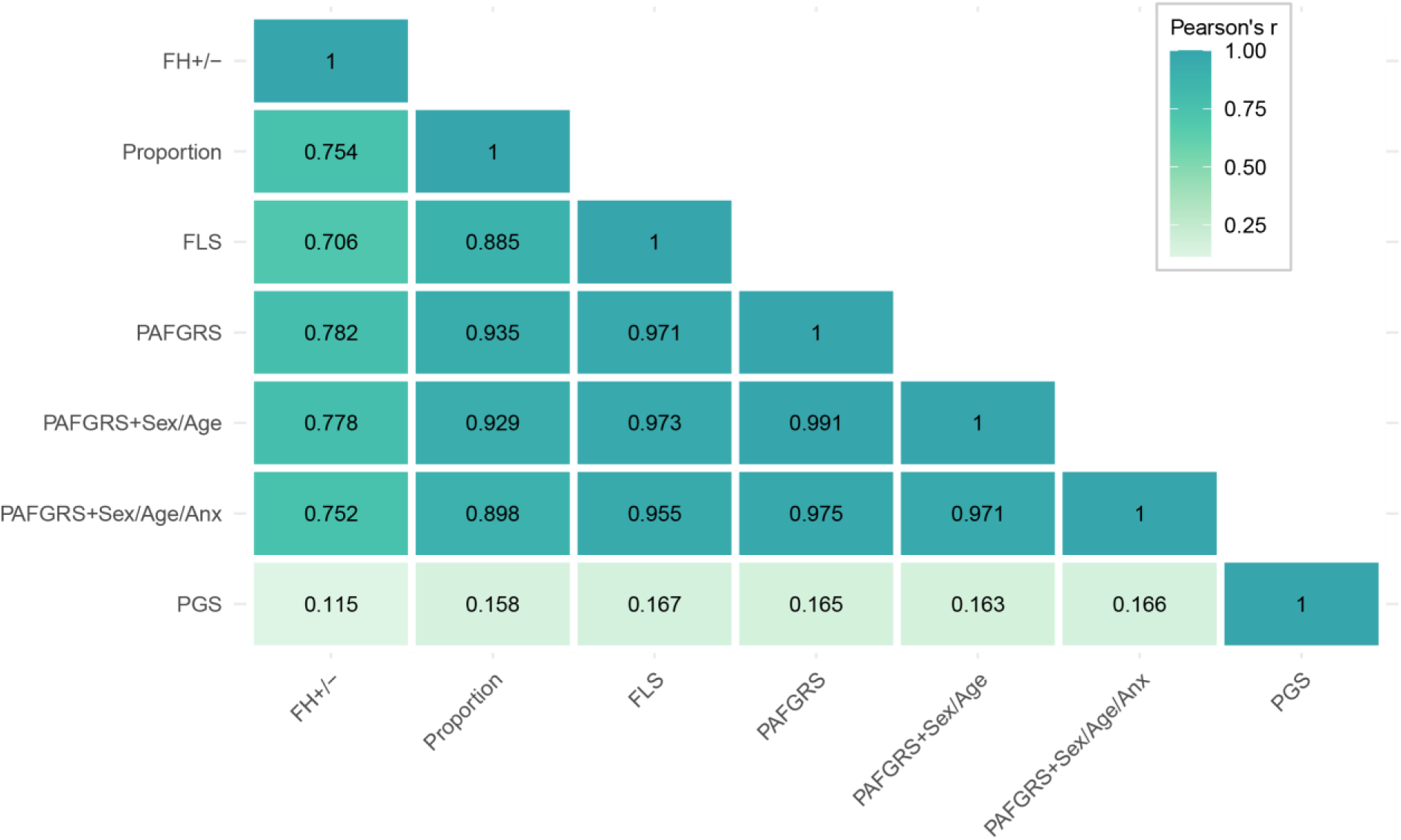
Pearson correlations between family history indicators and PGS. FH+/-: Family history of depression present/absent; FLS: Familial Liability Score; PAFGRS: Pearson-Aitkens Family Genetic Risk Score; ANX: Anxiety; PGS: Polygenic Score.

### Prediction of depressive disorder by family history indicators and PGS

Significant associations of genetic indicators with depressive disorder were confirmed by logistic regression models across predictors (**Figure 4a, Table S3**). Predictive results for extended assessment and imputation can be found in **Figure S4**. Overall, predictive accuracy was moderate, ranging from an AUC of 0.59 to 0.71. The predictive accuracy of the more complete family history indicators FLS (AUC = 0.68) and PAFGRS (AUC = 0.68, 0.69, 0.70) was slightly greater than that of the less complete indicators like FH=+/- (AUC = 0.62) and Proportion (AUC = 0.65), albeit with overlapping confidence intervals. As expected, predictive accuracy of the PGS was lower than that of all family history indicators (AUC = 0.59).

**Figure 4:**
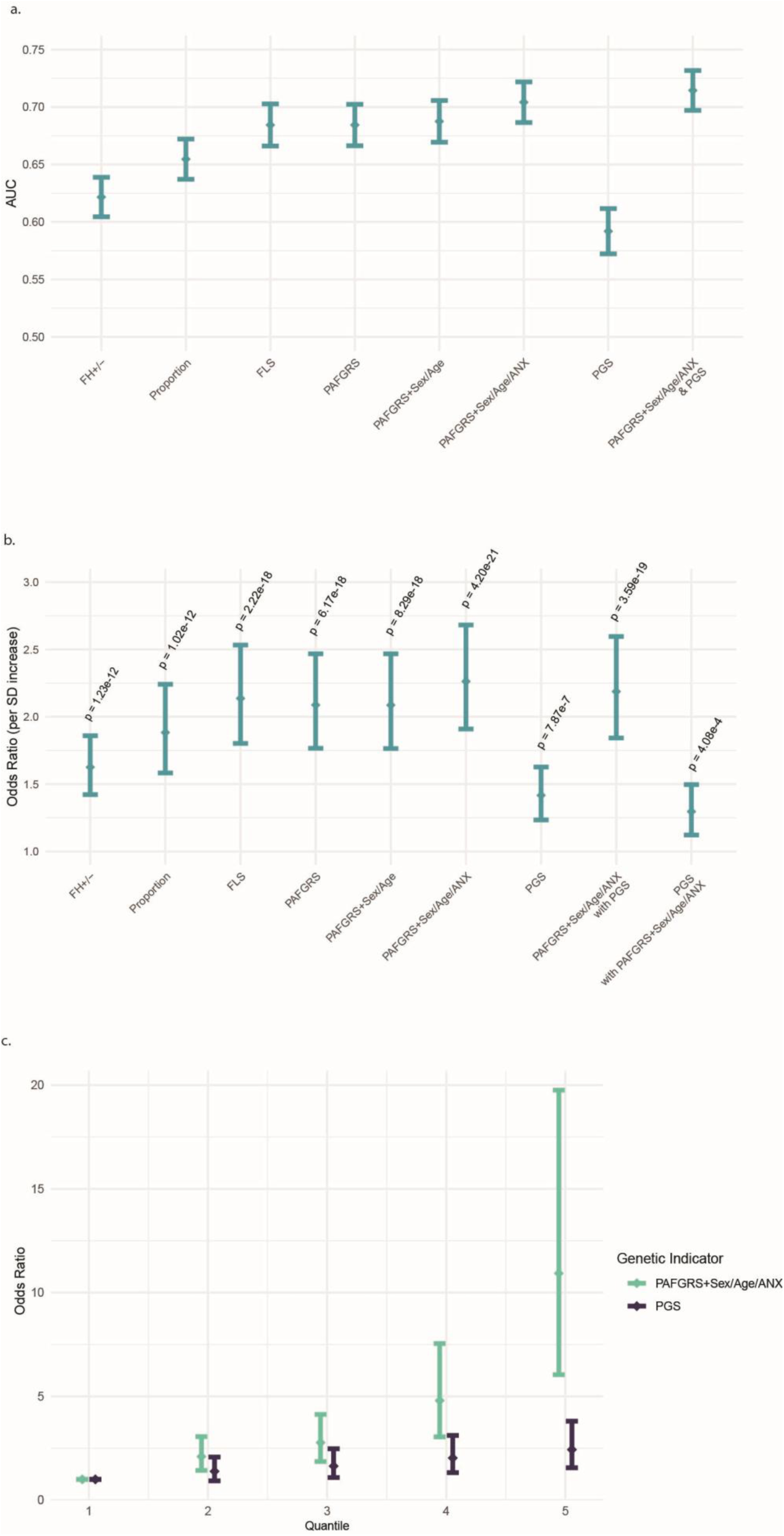
Results of logistic regression model predicting depressive disorder from genetic indicator(s). **PAFGRS: Pearson-Aitkens Family Genetic Risk Score; ANX: Anxiety; PGS: Polygenic Score.** **a. AUC of logistic regression model**. FH+/-: Family history of depression present/absent; FLS: Familial Liability Score; AUC: Area under the curve; P: p-value of the regression coefficient for the respective family history indicator; In combined model of family history and PGS, P(FH): p-value for PAFGRS+Sex/Age/ANX; P(PGS): p-value for PGS. For full results see **Table S3**. **b. Odds ratios of logistic regression model**. FH+/-: Family history of depression present/absent; FLS: Familial Liability Score; SD: standard deviation. Error bars represent 95% confidence intervals. For full results see **Table S3**. **c. Odds Ratios for depression in PAFGRS+Sex/Age/ANX and PGS by quintile**. Error bars represent 95% confidence intervals. Due to low sample size, we chose to divide the sample into quintiles; we note that this results in less pronounced odds ratios compared to dividing into deciles, as in e.g. Adams et al. 2025.

Adding the PGS to the best-performing family history indicator (PAFGRS+Sex/Age/ANX) slightly increased predictive accuracy (AUC = 0.71), albeit with overlapping confidence intervals. Regression coefficients were attenuated compared to a univariate model (**Table S3**), suggesting that family history and PGS are complementary in predicting depressive disorder.

Further detailing the relationship with depressive disorder, there was a clear trend that those in higher quantiles of family history and PGS have a higher risk for depression (**Figure 4c**). The family history indicator shows a stronger relationship with depressive disorder than the PGS, as expected from its greater AUC (**Figure 4a**). This was particularly noticeable in the fifth quantile, i.e. the 20% of persons with the greatest liability. The estimates in larger quantiles also show increasing confidence intervals, which is primarily due to non-linear scaling from the log odds scale to the OR scale, and partly due to the fifth quantile having a less balanced case control ratio (**Table S4**).

## Discussion

We directly compared a range of elaborate family history indicators to each other regarding their prediction of depressive disorder in a large clinical cohort study with established psychiatric diagnoses, and by including simple and convenient indicators such as dichotomous and proportion scores.

We found high correlations between the family history indicators, with a tendency for scores based on more types of input and more precise methods to be more predictive. Analysis alongside a polygenic score indicated that family history and genotypic data contain complementary information such that combining the two can improve prediction.

Patterns of associations between genetic indicators were in line with expectations and previous literature, providing important context for our prediction models. The correlations between continuous family history indicators were particularly high (>0.88). We thus extend previous findings of high correlations between elaborate family history indicators (Krebs et al. 2024) by highlighting that – in a clinical cohort with self-reported family history – a simpler continuous indicator—such as the proportion of family members affected – is also highly correlated with more complex scores.

More elaborate indicators were found to be slightly more predictive of depressive disorder, suggesting that added information on family size, sex and age of relatives, disorder prevalence and heritability, do capture relevant signal rather than noise.

The dichotomous indicator showed the lowest predictive accuracy, in line with previous findings (Milne et al. 2008). Differences between elaborate disorder-informed indicators were not significant, which may be attributable to the relatively small NESDA sample and assessment of first-degree relatives only.

When selecting a family history indicator for research or clinical practice, practical applicability of methods becomes important. Challenges faced by older non-genetic elaborate scores include (a) the treatment of missing data, (b) considering age at risk, (c) considering co-occurring disorders (Ali et al. 2021). All of these are solved by novel methods like PAFGRS, which offers a theoretically neat way of expanding the modelled liability with available data on a-c. The PAFGRS method also flaunts inherent usability and transferability across datasets. Overall, our results suggest that elaborate family history indicators are best suited to predict depression status, with PAFGRS additionally benefiting from the theoretical and practical advantages of modelling risk on the liability scale.

Across family history indicators, predictive accuracy was limited (AUC 0.62-0.70). However, even ideal genetic indicators are limited in explanatory power by the outcome trait’s heritability (depression: h²Twin = 0.35 [Polderman et al. 2015], h²SNP = 8.4% [Adams et al. 2025]) (Baselmans et al. 2021). Additionally, benchmarking family history indicators against the PGS highlighted the predictive value of family history.

The limited performance of the PGS may in part reflect lower predictive performance in NESDA (with *r_l_*^2^ = 0.037) compared to other studies (*r_l_*^2^ = 0.058 averaged across 42 European cohorts in Adams et al. [2025]). Nevertheless, PGS predictive accuracy (AUC 0.58) was in line with results from Krebs et al. (2024) (AUC 0.59), whereas PAFGRS predictive accuracy was slightly higher in our study (AUC 0.68-0.70) compared to the previous study (AUC 0.60-0.62). We hypothesize that this difference may be attributable to more severe and more accurately classified cases in the clinical NESDA cohort.

Both the divergence in predictive accuracy and the low correlations between family history indicators and PGS (0.11-0.16), are explained by the different aspects of genetic liability they capture. Firstly, PGS are still based exclusively on common genetic variants whose effects are inevitably estimated with noise. The precision of PGS in our study may for instance be affected by the genetic correlation between depression in the discovery GWAS (with assessment ranging from self-report to clinical diagnosis) and depression in NESDA being smaller than 1 (NESDA does not have the sample size to test this adequately). Secondly, even if SNP effects could have been assessed with full precision, the gap between SNP-heritability and family-based heritability has been well described (Matthews & Turkheimer 2022) and may be attributable to family history capturing other causes of genetic variation, such as rare variants, gene-gene interactions, or indirect genetic effects via parental genotypes. Third, inclusion of shared environmental effects (c² <= 0.1) in family history indicators can reduce its correlation with the PGS (Krebs et al. 2023). Although shared environmental effects are generally viewed as negligible in the field of behavioral genetics (Plomin et al. 2016), moderate influences can be detected for psychiatric phenotypes (Burt et al. 2009, Polderman et al. 2015).

In contrast to our results predicting depressive disorder (prevalence 0.2), lower predictive accuracy of family history is expected in rare disorders, as absence of affected family members in many families may lead to insufficient differentiation. Indeed, simulations have shown that family history indicators generally perform well in common disorders, whereas SNP-based scores perform better in rare diseases (So et al. 2011, Do et al. 2012).

Additionally, Krebs et al. (2024) demonstrated that the correlation between PAFGRS and PGS is a function of the modelled trait’s heritability, prevalence, and training GWAS used for the PGS. Training GWAS sample size (Adams et al. 2025, Howard et al. 2019) may be an explanation for the current results slightly exceeding correlations between PGS and PAFGRS observed in the iPsych Biobank (0.072-0.079, Krebs et al. 2023).

We found that although PAFGRS and PGS were both associated with depressive disorders, they still captured partially independent portions of disease liability, as indicated by the limited attenuation of their individual statistically significant associations when they were jointly modelled. This highlights the added value of including both family history and polygenic scores in prediction models. Simulations have shown that family history and SNP-based indicators are likely to remain complementary in the near future, with family history proving especially valuable in more common traits and when training GWAS for the PGS are smaller (Krebs et al. 2023).

Previous studies have successfully combined genotype and family history (albeit often dichotomous), for instance improving prediction of schizophrenia (Lu et al. 2017, Lu et al. 2022), and obesity (Wang et al. 2025). Family history data has also been included into molecular genetics studies to optimize the power of genome-wide association studies (Liu et al. 2017, Hujoel et al. 2020, Pedersen et al. 2022).

Comparison between family- and genotype-derived measures remains an area for further research investigating cutting-edge elaborate indicators and diverse disease phenotypes.

### Limitations

Our study is subject to a number of limitations. First, for comparing predictors with relatively small differences in prediction accuracy, the available sample had suboptimal power. However, the NESDA sample benefits from deep phenotyping with clinically validated depressive disorder assessment and detailed family history assessment. Second, the case-control ratio in NESDA is unbalanced (81% cases), as recruitment focused on patients with clinical depression or anxiety. However, the AUC metric is robust to oversampling of cases (Hand & Till 2001). Third, the family history data was based on retrospective self-report, potentially leading to recall bias, which may result in under-reporting by controls (Weissman 1987) and over-reporting by cases (Heun & Müller 1997). However, analysis of validation questions -- referring for instance to medication use – did not change results (**Figure S4**), suggesting that the impact of recall bias may have been limited. Importantly, self-report assessment is also reflective of realistic clinical settings in e.g. the Dutch healthcare system. Fourth, only first-degree relatives were included, which may yield less power compared to also including second- and third-degree relatives (Do et al. 2007). However, reports about second-degree relatives are expected to be less informative for prediction due to the smaller genetic relationship, may be subject to greater measurement error, and would be harder to assess in clinical practice. Fifth, age of onset in affected relatives is unknown in our sample, which may constitute a lack of information that could have been used to improve prediction. However, in the PAFGRS model its impact can to some extent be compensated by modelling age at interview.

## Conclusion

We conclude that elaborate family history indicators are better suited to predict depressive disorder. Our study extends previous findings to new data, specifically self-report in a clinical cohort which reflects a realistic clinical scenario. In addition, we showed that family history complements genotypic scores in the prediction of lifetime depression, providing insights relevant to both research and clinical practice.

## Data Availability

All summary-level data produced in the present study are available upon reasonable request to the authors. Individual-level NESDA data can be requested by researchers with an approved analysis plan, see https://www.nesda.nl/researchers/about-nesda/. The main scripts used for preprocessing and analysis of the data are available through GitHub.

https://github.com/EmmaPru/family-history-NESDA

## Disclosures

### Funding sources

EP, BWJHP, WJP and MB are supported by Stress in Action (NWO gravitation grant number 024.005.010). YM is supported by EU H2020 TO_AITION (grant number: 848146), Amsterdam UMC (Starter Grant Ronde 2), Amsterdam Neuroscience (PoC funding 2024-2026) and the ImmunoMIND consortium, funded by UK Research & Innovation as part of the UK national Mental Health Platform. The work of WJP was financially partly supported by the Amsterdam Cohort Hub, which is part of the Sector Plan ’Accelerating Health’ of the Dutch Ministry of Education, Culture and Science.

### Conflict of interest

The authors declare no biomedical financial interests or potential conflicts of interest.

## Acknowledgements

The infrastructure for the NESDA study (www.nesda.nl) is funded through the Geestkracht program of the Netherlands Organisation for Health Research and Development (ZonMw, grant number 10-000-1002) and financial contributions by participating universities and mental health care organizations (Amsterdam University Medical Centers (location VUmc), GGZ inGeest, Leiden University Medical Center, Leiden University, GGZ Rivierduinen, University Medical Center Groningen, University of Groningen, Lentis, GGZ Friesland, GGZ Drenthe, Rob Giel Onderzoekscentrum).

## Code availability

The main scripts used for preprocessing and analysis of the data are available through GitHub (https://github.com/EmmaPru/family-history-NESDA).

## CRediT author statement

Conceptualization, Methodology: EP, WP. Formal analysis: EP, PB. Data Curation: EP. Writing - Original Draft: EP, WP. Writing - Review & Editing: All authors. Funding acquisition: BWJHP.

## Notes

### Competing Interest Statement

The authors have declared no competing interest.

### Author Declarations

The Ethical Review Board of the VU University Medical Centre (Medisch Ethische Toetsingscommissie Vumc) gave ethical approval for the study protocol of NESDA.

