## Supplemental Material for "Comparing family history indicators and polygenic scores in depressive disorder"

### Supplemental Methods

#### Genotyping and imputation

Genotyping was done on two different platforms for the included NESDA participants, namely Perlegen-Affymetrix for 119 samples and on Affymetrix 6.0 for 2273 samples. On each platform, genotyping was performed following manufacturers protocols, using the then appropriate calling software. For each genotype platform, samples were removed if DNA sex did not match the expected phenotype, if the PLINK heterozygosity F statistic was < -0.10 or > 0.10, or if the genotyping call rate was < 0.90. SNPs were removed if the MAF < 1×10-6, if the Hardy-Weinberg equilibrium p-value was < 1×10-6, and/or if the call rate was < 0.95. Subsequently, for each platform, the genotype data was aligned with the 1000 Genomes reference panel using the HRC and 1000 Genomes checking tool, which tests and filters for SNPs with allele frequency differences larger than 0.20 as compared to the CEU population, palindromic SNPs and DNA strand issues.

Further processing took place on a combined dataset together with participants of the Netherlands Twin Register (NTR), genotyped on six different platforms. The NTR and NESDA data of the six platforms was then merged into a single dataset, keeping all quality controlled SNPs of each platform. For each individual, one platform was chosen. Based on the ~10.8k SNPs that all platforms have in common, DNA Identity By Descent state was estimated for all individual pairs using the Plink and King programs. These estimates were then compared to the expected familial relations, and samples were removed if these failed to fit. CEU population outliers, based on per platform 1000 Genomes PC projection with the Smartpca software, were removed from the data. Then, per platform, the data was phased using Eagle and then imputed to 1000 Genomes and Topmed using Minimac following the Michigan imputation server protocols. Post imputation, the resulting separate platform Variant Call Format (VCF) files were merged with Bcftools into a single VCF file per chromosome for each reference, only for those SNPs present on all six platforms.

#### Preprocessing of data for the PGS

Basic formatting and filtering steps were conducted with the help of PLINK (v2.0, Purcell and Chang 2019) and R (v4.3.2, R Core Team 2023). NESDA genome data was processed by removing SNPs with MAF < 1% or HWE < 0.0001, any occurrences of insertion or deletion, as well as duplicates. Any SNPs with imputation quality R² < 0.1 were also removed.

#### Processing of NESDA family histories: Simple and extended assessment of psychopathology (Example Depression)

##### Question Reference (English)

**G5**: Has your [relative] ever had mental problems such as depression, anxiety disorders, addiction, or other psychiatric problems?
**D1**: Has your [relative] ever had a depressive episode?
**D2**: Has your [relative] ever had an episode during which it seemed as if they were no longer interested in things they liked before, or an episode during which they could not enjoy things anymore?
**D5**: Was your [relative] ever treated for depression by a professional caregiver (GP, psychologist, psychiatrist)?
**D6**: Has your [relative] ever used medication for this?
**D7a**: Has your [relative] ever been admitted to an inpatient clinic for this?
**D7b**: During the inpatient treatment, did your [relative] undergo ECT?

##### General psychological

Questions were asked for each indicated first degree relative, one after the other (mother, father, siblings). If presence of any kind of psychological problems was established (**G5**), the interviewer moved on to the specific disorder questions for depression, anxiety, addiction, and other. For depression, follow-up questions (**D5**-**D7**) were asked only if the participant had indicated depressive symptoms in the first two core questions (yes to **D1** and/or **D2**).

##### Short assessment

The short assessment of depression psychopathology in relatives relied on two questions: **G5** and **D1**. If the answer was ‘yes’ on **D1** (which implies ‘yes’ on **G5**, otherwise the question would not have been asked), the relative is assigned depression status. If the answer was ‘no’ on either **G5** (no psychological problems, specific disorder questions skipped) or **D1** (no depressive episode), the relative was assigned control status. In all other cases (**D1** = ‘don’t know’/missing; **G5** = ‘yes’/‘don't know’) psychopathology status was set to missing.

##### Extended assessment

For the extended assessment of depression psychopathology, depression status was only assigned to the relative if symptoms indicated in the core questions (**D1** and/or **D2**) were confirmed by at least one of the follow-up questions (**D5**-**D7**).

Control status was assigned if either the answer to **G5** was ‘no’, or the answer to **D1** and/or **D2** was ‘no’, and the answer to neither **D1** nor **D2** was ‘yes’. Status was set to missing if both core items were missing, or if the answer to **D1** and/or **D2** was ‘yes’, but this indication of depressive symptoms was not confirmed by the follow-up questions.

##### Imputation

For both types of assessment, we additionally compared a) excluding missing data to b) imputation of relatives with missing data as controls. We decided to explore this simple way of imputing the data (rather than a more elaborate process based for instance on prevalences), based on the large proportion of unaffected persons in the population (80%) and potentially increased uncertainty in reporting on unaffected relatives.

### Supplemental Tables

##### Table S1: Comparison of short and extended family history assessment in NESDA.

***a****.* ***Cross-table short versus extended assessment of depression disorder in family members.*** *Number of relatives that were categorized as likely to be affected with depression (Case) or unaffected with depression (Control) or NA, under each assessment method (see Methods/Supplemental Methods).*

|  |  | **Extended assessment** | | |
| --- | --- | --- | --- | --- |
|  |  | **Control** | **Case** | **NA** |
| **Short assessment** | **Control** | 3468 | 33 | 72 |
|  | **Case** | 0 | 1040 | 464 |
|  | **NA** | 0 | 19 |  |

***b****.* ***Characteristics of short and extended assessment of depressive disorder in family members.*** *Contrasting short and extended assessment regarding basic data characteristics. Cases and Controls refer to NESDA participants themselves (as opposed to relatives). N: Number of NESDA participants; K: Prevalence; FH+: Family history of depression present; AUC: Area Under the Curve. PAFGRS: Pearson-Aitkens Family Genetic Risk Score; Dep.: Depressive Disorder; PRS: Polygenic Risk Score.*

|  | **Family history assessment** | |
| --- | --- | --- |
|  | **Short** | **Extended** |
| **N (Cases)** | 1059 | 1035 |
| **N (Controls)** | 250 | 250 |
| **K in relatives (Cases)** | 0.333 | 0.268 |
| **K in relatives (Controls)** | 0.153 | 0.137 |
| **FH + (Cases)** | 0.686 | 0.576 |
| **FH + (Controls)** | 0.443 | 0.395 |
| **AUC of PAFGRS (Dep. prediction)** | 0.684 | 0.652 |
| **Correlation PAFGRS-PRS (Pearson's)** | 0.165 | 0.167 |

##### Table S2. Effect of imputation on number of relatives.

**a. Number of relatives and prevalence of psychopathology amongst relatives.** Gain in number of relatives (compare N and N [imputed]) is substantial (up to 1821 relatives). K: Prevalence of psychopathology; N: Sample size.

|  | **K** | **K (imputed)** | **N** | **N (imputed)** |
| --- | --- | --- | --- | --- |
| **Depression** | | | | |
| **Short** | 0.296 | 0.250 | 5077 | 6026 |
| **Extended** | 0.239 | 0.181 | 4560 | 6026 |
| **Anxiety** | | | | |
| **Short** | 0.142 | 0.119 | 5073 | 6026 |
| **Extended** | 0.137 | 0.103 | 4519 | 6026 |

***b. Cross-table of NESDA participant sample size****. Gain in number of index participants is small (up to n = 54).*

|  | **Basic** | **Imputation** |
| --- | --- | --- |
| **Short assessment** | 1309 | 1339 |
| **Extended assessment** | 1285 | 1339 |

##### Table S3. Detailed prediction results.

ROC: Receiver operator curve; FH+/-: Family history of depression present/absent; FLS: Familial Liability Score; PAFGRS: Pearson-Aitkens Family Genetic Risk Score; ANX: Anxiety; PRS: Polygenic Risk Score. AUC: Area under the curve; OR: Odd's ratio; SE: Standard error; 95% CI Lower: lower limit of 95% confidence interval; 95% CI Upper: upper limit of 95% confidence interval.

|  | **ROC summary** | | | **Logistic regression** | | | | | |
| --- | --- | --- | --- | --- | --- | --- | --- | --- | --- |
| **Predictor** | **AUC** | **OR** | **SE (AUC)** | | **Coefficient** | **SE (Coefficient)** | **95% CI Lower** | **95% CI Upper** | **P-value** |
| FH+/- | 0.622 | 1.63 | | 0.017 | 0.486 | 0.068 | 1.42 | 1.86 | 1.23E-12 |
| Proportion | 0.655 | 1.88 | | 0.018 | 0.633 | 0.089 | 1.58 | 2.24 | 1.02E-12 |
| FLS | 0.684 | 2.14 | | 0.018 | 0.759 | 0.087 | 1.8 | 2.53 | 2.22E-18 |
| PAFGRS | 0.684 | 2.09 | | 0.018 | 0.736 | 0.085 | 1.77 | 2.47 | 6.17E-18 |
| PAFGRS+Sex/Age | 0.688 | 2.09 | | 0.018 | 0.736 | 0.086 | 1.76 | 2.47 | 8.29E-18 |
| PAFGRS+Sex/Age/ANX | 0.704 | 2.26 | | 0.018 | 0.817 | 0.087 | 1.91 | 2.68 | 4.20E-21 |
| PRS | 0.592 | 1.42 | | 0.02 | 0.348 | 0.071 | 1.23 | 1.63 | 7.87E-07 |
| PAFGRS+Sex/Age/ANX (covariate PRS) | 0.714 | 2.19 | | 0.017 | 0.783 | 0.087 | 1.84 | 2.6 | 3.59E-19 |
| PRS (covariate PAFGRS+Sex/Age/ANX) | 0.714 | 1.3 | | 0.017 | 0.259 | 0.073 | 1.12 | 1.5 | 0.000408449 |

##### Table S4. Detailed results for logistic regression per quintile.

SE: Standard error on the log odds scale before transformation to odd's ratios confidence intervals; N: Number of participants in respective quintile. PAFGRS: Pearson-Aitkens Family Genetic Risk Score; ANX: Anxiety; PRS: Polygenic Risk Score.

|  | **PAFGRS+Sex/Age/ANX** | | | **PRS** | | |
| --- | --- | --- | --- | --- | --- | --- |
| **Quantile** | **SE** | **N (Cases)** | **N (Controls)** | **SE** | **N (Cases)** | **N (Controls)** |
| **1** | 0.126 | 167 | 101 | 0.138 | 196 | 72 |
| **2** | 0.193 | 208 | 60 | 0.204 | 212 | 56 |
| **3** | 0.203 | 220 | 48 | 0.210 | 219 | 49 |
| **4** | 0.231 | 238 | 30 | 0.219 | 227 | 41 |
| **5** | 0.302 | 253 | 14 | 0.228 | 232 | 35 |

### Supplemental Figures

**Figure S1:** **Family history interview questions with illustration of short versus extended assessment.**

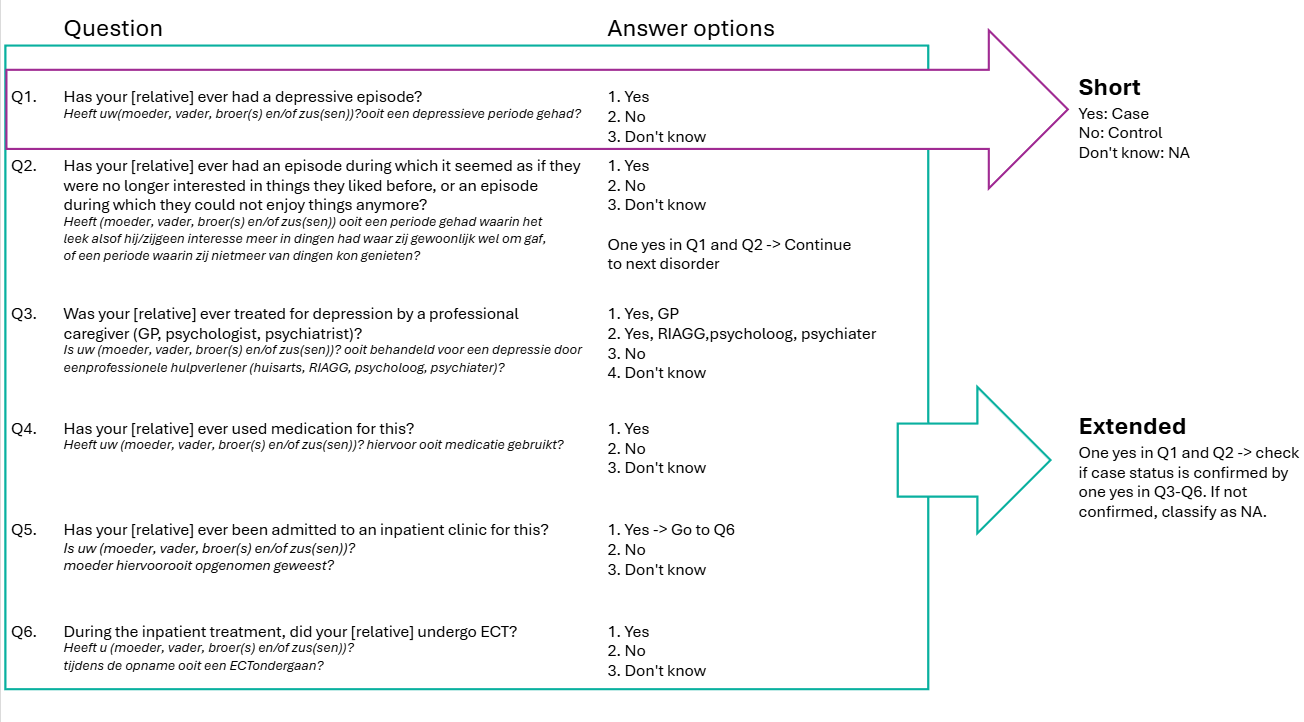

##### Figure S2: Input for PAFGRS+Sex/Age/ANX: Age- and sex-specific cumulative incidences.

Values based on McGrath et al. 2023 and scaled for the Netherlands based on de Graaf et al. 2012. PAFGRS: Pearson-Aitkens Family Genetic Risk Score; ANX: Anxiety.

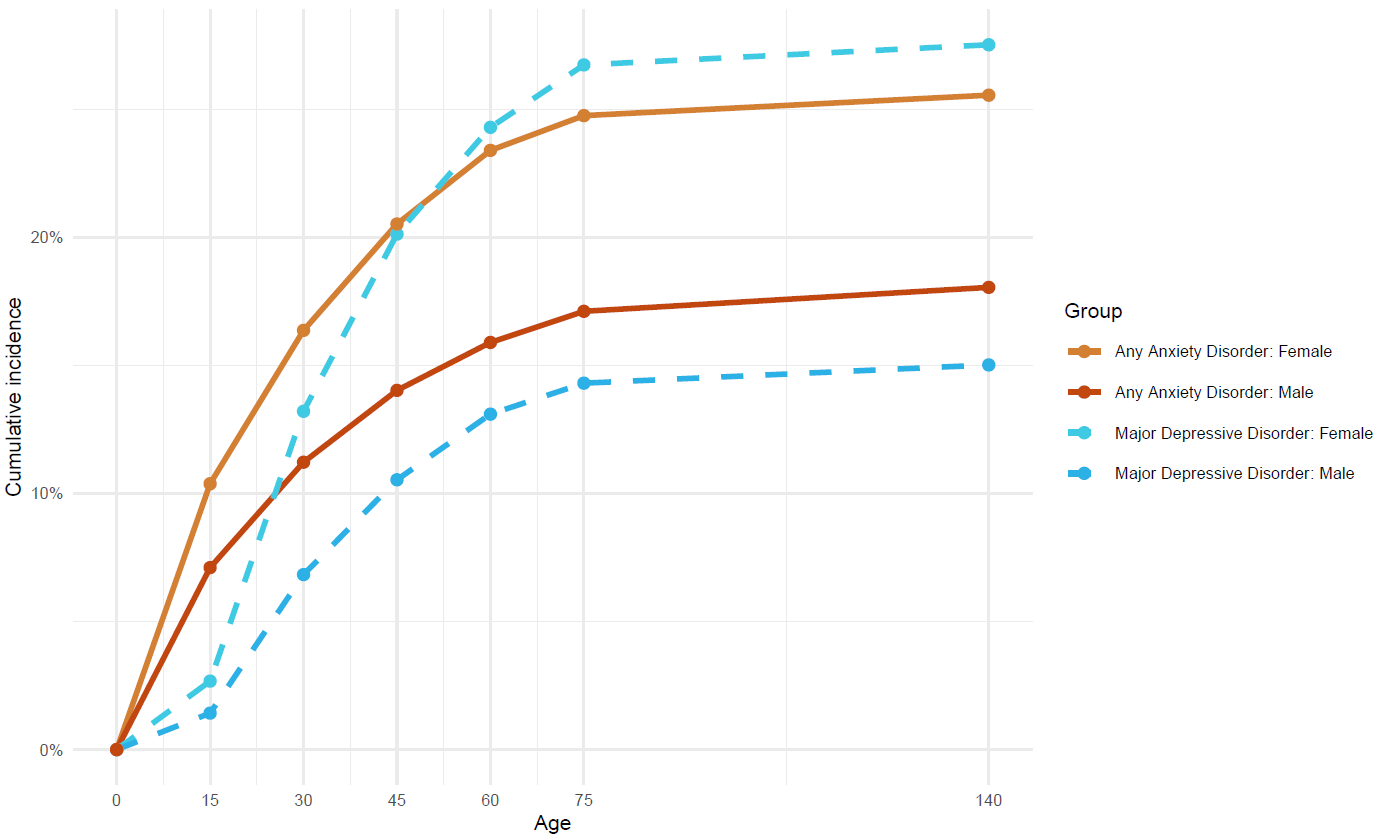

##### Figure S3: Spearman's rank correlations between family history indicators and PRS.

FH+/-: Family history of depression present/absent; FLS: Familial Liability Score; PAFGRS: Pearson-Aitkens Family Genetic Risk Score; ANX: Anxiety; PRS: Polygenic Risk Score.

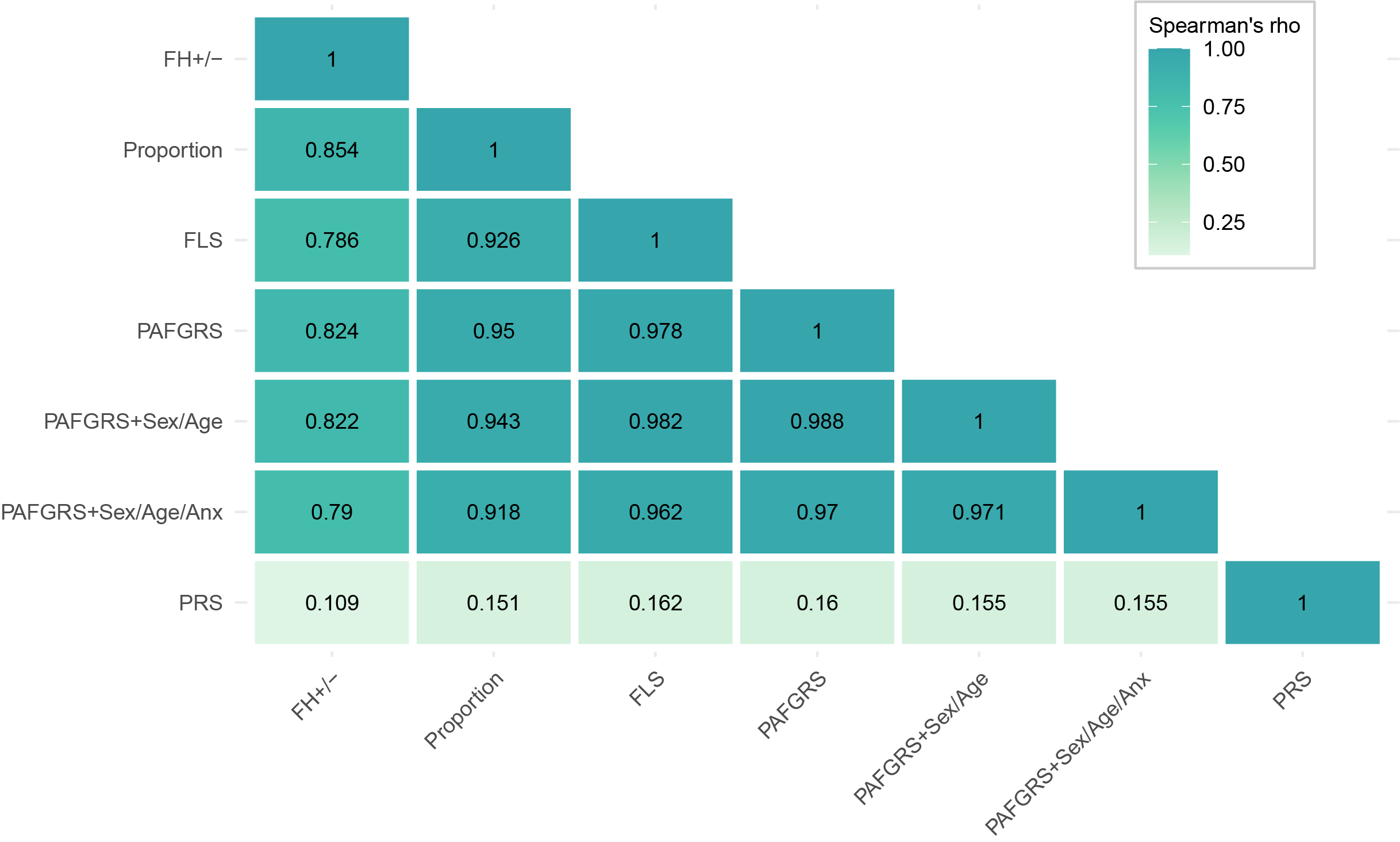

##### Figure S4: AUC of logistic regression model predicting depression status from genetic indicator(s).

Extended family history assessment and imputation. FH+/-: Family history of depression present/absent; FLS: Familial Liability Score; PAFGRS: Pearson-Aitkens Family Genetic Risk Score; ANX: Anxiety; PRS: Polygenic Risk Score.

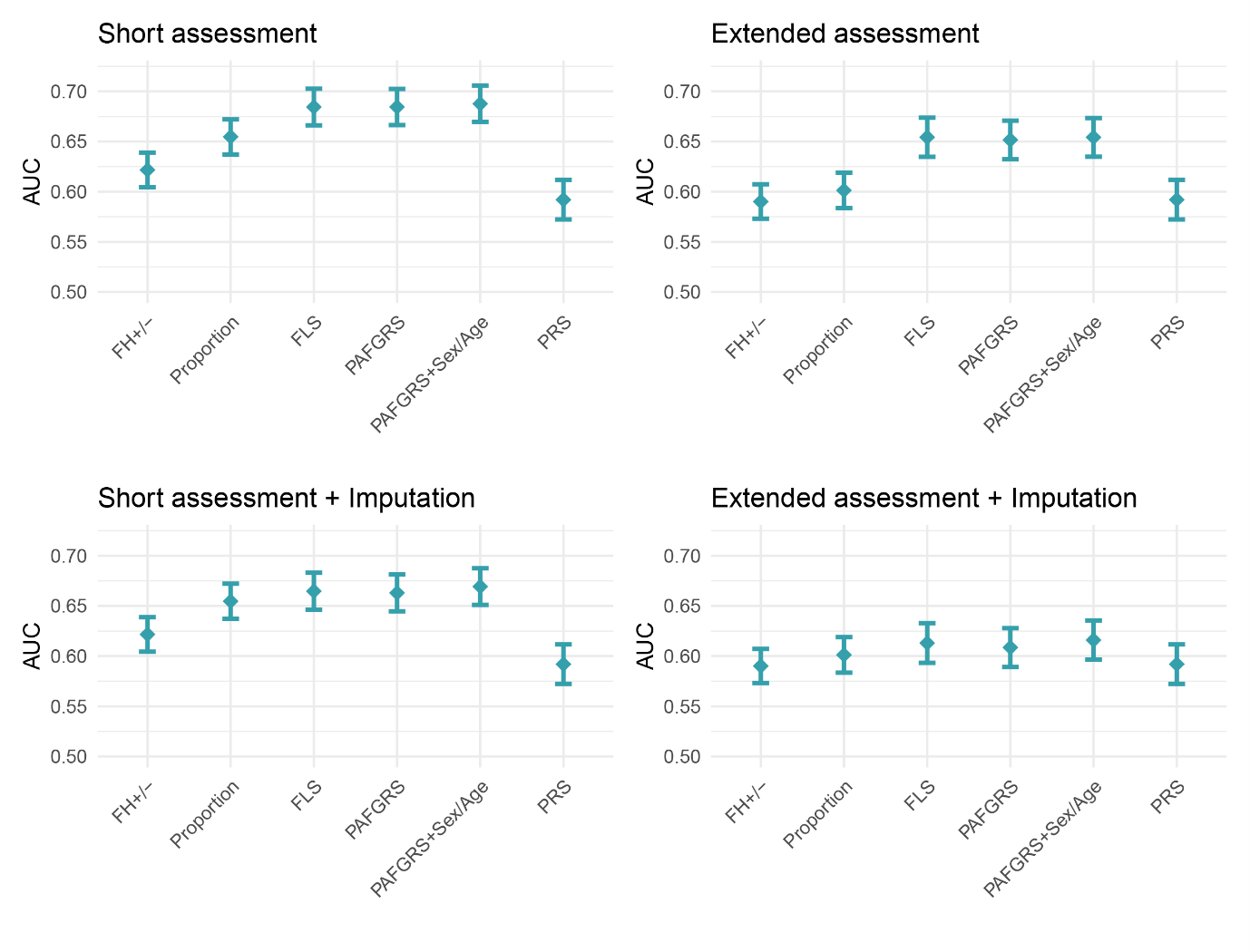
